# Concurrent Transcranial Magnetic Stimulation and Electroencephalography Measures are Associated with Antidepressant Response from rTMS Treatment for Depression

**DOI:** 10.1101/2023.02.10.23285794

**Authors:** Neil W. Bailey, Kate E. Hoy, Caley M. Sullivan, Brienna Allman, Nigel C. Rogasch, Zafiris J. Daskalakis, Paul B Fitzgerald

## Abstract

**Background:** Response rates to repetitive transcranial magnetic stimulation (rTMS) for depression are 25-45%. Biomarkers predicting response to rTMS may reduce treatment burden. TMS-evoked neural activity recorded via electroencephalography (EEG) has potential as a biomarker of treatment response. We examined whether these measures could differentiate responders and non-responders to rTMS for depression.

**Methods:** Thirty-nine patients with treatment-resistant major depressive disorder (MDD) and 21 healthy controls received TMS during EEG recordings (TMS-EEG). MDD participants then completed 5-8 weeks of rTMS treatment. Repeated measures ANOVAs compared N100 amplitude, N100 slope, and theta power across 3 groups (responders, non-responders and controls), 2 hemispheres (left, F3, and right, F4), and 2 stimulation types (single pulse and paired pulses with a 100ms inter-pulse interval [pp100]).

**Results:** Neither N100 amplitude nor theta power differed between responders and non-responders. The control group showed more negative N100 amplitudes than the combined depression group. Responders showed a steeper negative N100 slope for single pulses and steeper positive slope for pp100 pulses at F3 than non-responders. Exploratory analyses suggested this may have been due to the responder group showing larger late P60 and N100 amplitudes. Receiver Operator Characteristic (ROC) curve analysis indicated the difference between single and pp100 slopes provided excellent sensitivity (1.00), but poor specificity (0.455).

**Limitations:** Our study had a small sample size.

**Conclusion:** Left hemisphere TEPs, in particular N100 slope, may be useful in predicting non-responders to rTMS treatment for depression. Non-response prediction may be useful in saving clinical resources and patient time.

## 1. Introduction

Treatment resistance is common in major depression, with estimates suggesting that up to 50% of people with depression fail to respond to at least one antidepressant treatment (Nemeroff, 2007). Repetitive transcranial magnetic stimulation (rTMS) has become a common alternative when antidepressant medications do not work. However, meta-analyses have suggested response rates to rTMS range between 25-45% (Berlim et al., 2013, Cao et al. 2018, Sehatzadeh et al. 2019). Despite these response rates, patients who do respond to rTMS often show considerable improvements, while non-responders often show very little change in depression severity, such that response rates demonstrate a bimodal distribution (Fitzgerald et al., 2016). This highlights the potential for patients to be categorized as responders or non-responders based on physiological markers of response likelihood obtained prior to treatment onset. In an attempt to alleviate the significant costs to clinics and burdens to patients in unnecessary cases when patients will fall into the category of ‘non-responder’, research has recently begun to focus on prediction of treatment response (Kar, 2019). Achieving this goal could be a first step towards individualised treatments for depression, so that a prediction of non-response to one treatment could indicate use of another treatment, leading to more rapid and successful depression treatment responses.

A growing amount of research using both electroencephalography (EEG) and functional magnetic resonance imaging (fMRI) measures has achieved varying degrees of accuracy in prediction of response to rTMS (for review, see Kar (2019)). Some of the more successful examples of this research have combined neuroimaging with measures of early response one week following the start of treatment. Indeed, early change in depression rating scales appears to be one of the strongest predictors (Bailey et al. 2018, 2019). However, using measures of change in the first week of treatment is unlikely to have as much utility as truly predictive measures based on pre-treatment assessment only. Similarly, although fMRI measures have sometimes been highly accurate at predicting responders (Cash et al. 2019, Drysdale et al., 2017, however, also see Dinga et al., 2019), fMRI is expensive, and as such fMRI measures may be clinically impractical for response prediction. There are also potential issues with the reliability of fMRI measures, which may preclude their use as predictors of treatment response (see Elliott et al. 2020). Additionally, EEG and fMRI measures might measure biomarkers that are only indirectly specific to the underlying mechanisms that respond to rTMS (i.e., aspects of ongoing neural activity targeted by the stimulation). In contrast, measuring the neural response to a TMS pulse may provide a more sensitive predictor of treatment response due to the more direct analogue of the response to a TMS pulse to the effect of treatment - neural activity measured following a TMS pulse and rTMS treatments are governed by similar processes (Daskalakis and Tyndale, 2019).

Combining transcranial magnetic stimulation with EEG (TMS-EEG) has become a common research protocol and provides a measure of neural activity in response to TMS (Tremblay et al. 2019). These responses are known as TMS evoked potentials (TEPs) and TMS evoked oscillations. TEPs and TMS evoked oscillations are sensitive to both the neural responses following transcranial stimulation of the targeted cortical region (Gosseries et al. 2015) and also to sensory-evoked responses following auditory and somatosensory inputs associated with the clicking sound and sensation of TMS (Nikouline et al. 1999, Conde et al. 2019, Biabani et al. 2019, Biabani et al. 2021). The amplitude of some TEPs is sensitive to gamma-aminobutyric acid-B (GABAb) receptor inhibitory function - in particular the N100, a negative scalp recorded voltage deflection around 100ms following the TMS pulse (Premoli et al. 2014a, Premoli et al. 2014b, Rogasch et al. 2013, Rogasch et al. 2015, Cash et al. 2017, Belardinelli et al. 2021, Kaarre et al. 2018, Noda 2020). While there is evidence from studies of TMS applied to the motor cortex suggesting the N100 reflects a neural response to the TMS pulse (Rocchi et al. 2021), recent studies have shown that the N100 is also sensitive to sensory activity, particularly in frontocentral electrodes (Biabani et al. 2019, Conde et al. 2019, Gordon et al. 2018, Fernandez et al. 2021). Although this research suggests that the cause of potential differences in the N100 in responders compared to non-responders to TMS might be ambiguous (with differences due to either responses to sensory stimulation, responses to the TMS pulse, or both) the differences are still physiologically meaningful either way, and could offer predictive potential. Indeed, the N100 following stimulation of the left DLPFC has been found to be increased in people with depression compared to healthy individuals (Voineskos et al. 2019) and reduced with rTMS treatment (Voineskos et al. 2021). Furthermore, the size of the TEP before treatment is predictive of larger reductions in suicidal ideation following rTMS (Voineskos et al. 2021), as is a more negative N100 amplitudes prior to magnetic seizure therapy (Sun et al. 2018).

TEPs are often produced by single TMS pulses, but can also be modulated by paired pulse paradigms which deliver two TMS pulses in quick succession (e.g., 100 ms apart). The first of the paired pulses is thought to result in a period of cortical inhibition lasting ∼150-200 ms, so that when the second pulse is delivered 100 ms following the first, the TEP responses to the second pulse are reduced (Rogasch et al. 2013). Suppression of TEPs following paradigms using paired pulses separated by ∼100 ms (pp100) are thought to reflect GABAb receptor-mediated inhibitory mechanisms (Rogasch et al., 2014, 2015, Premoli et al. 2014b), although this relationship is likely complex (Ilmoniemi et al. 2021). The function of the GABAb receptor in the prefrontal cortex is important for memory, with both activity in the DLPFC being related to working memory performance, and TEP suppression following pp100 over the DLPFC being related to working memory performance (Goldman-Rakic, 1995; Gonzalez-Burgos et al., 2011; Jacobson et al., 2007; Michels et al., 2012; Mohler, 2009, Daskalakis et al., 2008b; Hoppenbrouwers et al., 2013; Rogasch et al., 2015, Miller and Cohen, 2001; Owen et al., 2005). In addition to relationships with memory, higher TEP suppression following pp100 prior to treatment is also predictive of reductions in suicidal ideation following magnetic seizure therapy (Sun et al. 2018).

Neural oscillations are also elicited by the TMS pulse and these TMS evoked oscillations can also be measured by TMS-EEG. The phase of EEG voltage shifts in response to the TMS pulse appears to be significantly time locked to the TMS pulse (Ferrarelli et al., 2008; Paus et al., 2001). The peak frequencies of oscillations that are elicited by TMS are also unrelated to the intensity of the TMS pulse, and the frequencies generated differ across different cortical regions (Fuggetta et al., 2005; Paus et al., 2001; Rosanova et al., 2009). People with depression show increased theta (4-7 Hz) TMS-evoked oscillations relative to healthy individuals prior to treatment, and show reductions in theta oscillations following electroconvulsive and magnetic seizure therapy (Hill et al. 2021). Furthermore, previous research has shown that theta activity related to working memory function predicted response to rTMS treatment (Bailey et al. 2018). Taken together, these findings suggest that both evoked and oscillatory activity following single TMS pulses are increased in people with depression, are altered by brain stimulation therapies, and may predict therapeutic response outcome.

Given this background, the aim of this study was to assess whether TMS-evoked neural activity following single and paired pulses differentiated responders and non-responders to rTMS treatment in people with depression. Following research that indicated N100 amplitude predicts reductions in suicide ideation from magnetic seizure therapy (Sun et al. 2018) we hypothesized that larger N100 amplitudes following single pulses, and more TEP suppression following pp100 will be present at baseline in individuals who will respond to rTMS treatment for depression. Additionally, following our previous research showing higher theta predicted non-response in working memory related EEG (Bailey et al. 2018), we hypothesized that TMS evoked theta oscillations would be larger in amplitude in responders to rTMS treatment for depression.

## 2. Methods

### 2.1 Participants

Fifty participants with treatment resistant major depressive disorder (MDD) and 21 healthy controls took part in the study. This study was part of a larger study of clinical responses to different rTMS treatment protocols (Fitzgerald et al. 2018), where half of the participants took part in a baseline and endpoint EEG and TMS-EEG assessments, and half of the participants took part in a baseline MRI scans, which has been published separately (Cash et al. 2019a, 2019b). Healthy controls participated in a single TMS-EEG session, while MDD participants completed a baseline TMS-EEG and endpoint TMS-EEG session after 5-8 weeks of rTMS treatment. Eight participants with MDD were excluded from analysis due to withdrawing from the study after one week of treatment (one due to headache/pain, one due to excessive travel time required for the trial, and six without providing a reason). Three further participants with MDD were excluded due to extremely noisy TMS-EEG data. Of the remaining 39 MDD participants, 11 were classified as responders to rTMS treatment, with reductions in the 17-item Hamilton Rating Scale for Depression (HRSD) (Williams, 1988) score of > 50%. For our endpoint recordings, a further one responder participant and six non-responders did not return for their final TMS-EEG session. The participants included in this study overlap with those reported previously in an analysis of resting and working memory related EEG (Bailey et al. 2018, 2019), and further demographic details and inclusion/exclusion criteria can be found in those publications. Ethical approval for the study was obtained from the Alfred Hospital and Monash University’s ethics committees, and all participants gave written informed consent before participating in the study.

### 2.2 Procedure

The procedure for the rTMS treatment and testing sessions is identical to that reported in Bailey et al. (2018). Participants with MDD underwent a baseline interview where demographic and depression severity data were collected, including the HRSD, Montgomery Asberg Depression Rating Scale (MADRS) (Montgomery and Asberg, 1979) and the Beck Depression Inventory-II (BDI-II) (Beck et al. 1996). All MDD participants underwent three weeks of week daily unilateral left 10Hz rTMS treatment. Individuals who showed a response to this treatment at week 3 were continued on an additional two weeks of titrated rTMS treatment (three sessions in week 4, two sessions in week 5), after which they completed another TMS-EEG session. Non-responders after three weeks were randomised to either continue unilateral left 10Hz rTMS treatment for another three weeks, or to unilateral right 1Hz rTMS, or sequential bilateral rTMS consisting of 1Hz right rTMS followed by 10Hz left rTMS treatment. Individuals who had not responded by week 3 but responded by week 6 were continued on an additional two weeks of titrated rTMS treatment in the same manner as those who had responded at week Left sided treatments were given to the F3 electrode location, while right sided treatments were given to the F4 electrode location. Further details can be found in Bailey et al. (2018).

### 2.3 TMS-EEG recordings and pre-processing

TMS-EEG recordings were performed in a darkened room using a 30 channel Ag/AgCl electrode EasyCap (EasyCap, Woerthsee-Etterschlag, Germany) to record EEG activity to Neuroscan Acquire software using a Synamps 2 amplifier (Compumedics, Melbourne, Australia). Electrodes used were AF3, AF4, F5, F3, F1, Fz, F2, F4, F6, FC5, FC3, FC1, FCz, FC2, FC4, FC6, Cz, P7, P5, P3, P1, Pz, P2, P4, P6, P8, PO3, PO4, O1, and O2, plus electrodes above and below the left eye and outside the outer rim of each eye to record eye movements. CPz was used as the online reference and AFz as the ground. Impedances of < 5 kΩ were maintained throughout the session. EEG was sampled at 10,000 Hz (with a DC high pass and 2,000 Hz low pass filter).

The EEG cap was applied, then the resting motor threshold (rMT) was determined applying TMS through the EEG cap. rMT was defined as the minimum TMS intensity required to elicit an average of 1mV response in the relaxed adductor pollicis brevis muscle from 10 TMS pulses (Naim-Feil et al. 2016; Zeimann et al. 1998). During the TMS-EEG recordings, participants listened to white noise through intra-auricular earphones (Etymotic Research, ER3-14A, USA) to limit the influence of the TMS click sound on the EEG signal (ter Braack et al. 2013). The volume for this white noise was set prior to the TMS-EEG. The white noise was played to the participants through these earphones, and the volume was adjusted to the maximum level they were comfortable with, which each participant confirmed masked the sound of the TMS pulse.

Both single and paired TMS pulses in the pp100 paradigm (with 100ms separating the two pp100 pulses) were delivered using a figure-of-eight MagVenture B-65 fluid-cooled coil (MagVenture A/S, Denmark) in biphasic mode. Seventy-five single and 75 pp100 pulses were delivered to each stimulation target in a randomized sequence, with an inter-trial interval of 5s +/- a 10% jitter at the 1mV rMT TMS intensity level. TMS pulses were applied to both the left hemisphere (with the TMS coil placed at a location between F3 and FC3) and right hemisphere (with the TMS coil placed at a location between F4 and FC4) with the coil positioned at a 45 degree angle relative to the midline, with the handle pointing posteriorly. Participants were randomised in equal numbers to receive left hemisphere or right hemisphere TMS first. Coil location was maintained using a stand, and the edge of the coil was marked on the cap for consistent re-positioning of the coil to compensate for any participant movements during the delivery of the pulses. This method has been demonstrated to be accurate to within 5mm when neuronavigation is not available (Rogasch, Thomson, Daskalakis, and Fitzgerald, 2013).

TMS-EEG data was processed using EEGLAB (Delorme and Makeig, 2004) and TESA (Rogasch et al. 2017) in MATLAB (2015b, The MathWorks, USA). First, EEG data were epoched around the TMS pulses (time locked to the TMS pulse for the single pulses, and to the second TMS pulse for the pp100 trials) from -1000ms to 1000ms and baseline corrected (-500 to -50ms). The period surrounding the large artefact generated by the TMS pulse was removed and interpolated (-5 to 15ms). Data were then downsampled to 1000Hz. Epochs were visually inspected and electrodes showing signals suggesting poor contact were removed from the entire dataset, as were epochs showing excessive noise. We then used a first round of ICA (FastICA with “tanh” contrast) to remove the muscle artefact following the pulse from the data using a semi-automated component classification algorithm, which identified the artefact if the component was > 8x larger than the mean absolute amplitude across the entire time course (tesa_compselect, Rogasch et al. 2017). Data were then band-pass filtered from 1-80Hz and band-stop filtered from 47-53Hz using a butterworth second order zero-phase filter. A second round of ICA was then performed to remove other non-neural artefacts, again using tesa_compselect which identified components as artefacts including eye movements (mean absolute z score of AF3 and AF4 > 2.5), persistent muscle activity (high frequency power >60% of total power), decay artefact and electrode noise (absolute z score of electrodes > 4). Excluded electrodes were then interpolated back into the data using spherical interpolation, data were re-referenced to the common average, and single pulse trials and pp100 trials were averaged separately in preparation for submission to our primary TEP analysis. Additionally, in an exploratory analysis to assess potential differences between groups in pp100 related suppression of the N100, a measure of pp100 related N100 suppression was calculated as follows: 1) pp100 TEPs were corrected for the influence of the conditioning pulse by subtracting averaged single pulse TEP data from the period that was temporally aligned with the pp100 conditioning pulse from the averaged pp100 TEP data, then 2) averaged single pulse TEP data temporally aligned with the pp100 test pulse was also subtracted from the pp100 data. This provided a measure of the difference between response to the single pulse and response to the pp100 test pulse, after controlling for the ongoing activity produced by the conditioning pulse, following the method provided by previous research (Opie et al. 2018). This approach results in a measure of the inhibitory influence of the conditioning pulse on the test pulse, without results being confounded simply by the voltage shifts produced by the conditioning pulse (Opie et al. 2018). This TEP data is referred to as pp100-corrected in our analyses.

### 2.4 TMS Evoked Potentials and Oscillations

We measured the N100 amplitude from F3 while stimulation was provided over F3, and F4 while stimulation was provided over F4, as the average amplitude from 85ms to 140ms after the TMS pulse (Rogasch et al. 2014, 2015, Gordon et al. 2018). The N100 slope is also correlated with measures of TEP suppression (Rogasch et al. 2013, 2015), so we measured N100 slope as the mean first derivative of the voltage from 90 to 98ms after the TMS pulse from F3 and F4 with left and right sided stimulation respectively (Rogasch et al. 2015).

Theta power was computed using a Morlet wavelet transform (3.5 cycles with 0.5 Hz steps), and was baseline corrected to a pre-stimulus period (-500 to -100 ms) using the relative method (active period – mean baseline period / mean baseline period) based on previous studies of TMS-evoked oscillations over DLPFC (Rogasch et al., 2014, Rogasch et al., 2015, Hill et al., 2017, Chung et al. 2017). Theta power was averaged across all trials, and across a window from 50 ms to 250 ms post stimulus, across the 4 to 8 Hz frequency band, and extracted from F3 while stimulation was provided over F3 and F4 while stimulation was provided over F4.

Separate 3 x 2 x 2 repeated measures ANOVAs were conducted for N100 amplitude and N100 slope, which compared the 3 groups (responders, non-responders and controls), 2 hemispheres (left, F3, and right, F4), and 2 stimulation types (single pulse and pp100). A 3 x 2 repeated measures ANOVA was conducted for pp100-corrected N100 data, comparing the 3 groups (responders, non-responders and controls) by stimulation at 2 hemispheres (left and right DLPFC). A 3 x 2 repeated measures ANOVA was conducted for theta power, comparing the 3 groups (responders, non-responders and controls) by stimulation at 2 hemispheres (left and right DLPFC). Additionally, to assess whether these TMS-EEG measures were altered by the TMS treatment, the same repeated measures ANOVAs were conducted from data obtained from the depression groups at the endpoint of treatment (END) and the baseline (BL) control group data (since the control group did not undergo TMS treatment and as such only had a baseline TMS-EEG recording). Post hoc comparisons with reduced statistical designs were planned to explore group main effects and potential interactions involving group, with statistical designs restricted to two groups only where differences between the responder, non-responder and control groups were present.

Data met assumptions for normality and skewedness (values of less than 2), with the exception of 3 N100 amplitude variables that exceed 2 on the Kurtosis measure. To address the violation of normality, an absolute value of 50 was added to all single pulse and pp100 N100 amplitude values, then values were log transformed to normalise the data and statistics were conducted based on these transformed values. Other variables were left as they were without transforms. Additionally, prior to analysis seven of the single pulse or pp100 N100 amplitude variables had a single outlier (z-score > 3.29) winsorized, 9 of the pp100-corrected N100 amplitude values were winsorized, one N100 slope variable had a single outlier winsorized, and each theta power variables had a single outlier winsorized.

Multiple comparison controls were implemented on main effects and interactions of interest from primary hypotheses using the false discovery rate (FDR) method (Benjamini and Hochberg, 1995). To enable comparison with future research, both corrected and uncorrected p-values are reported (labelled “FDR-p” and “p” respectively). Finally, because both inspection of the TEP figure and our N100 slope results suggested an effect that was not captured by our a priori selected window of interest, we used permutation statistics to perform an exploratory test of potential differences between the responder, non-responder and control groups across all timepoints within the single pulse condition applied to the left hemisphere. This analysis involved shuffling group labels without replacement 5000 times to produce a null result distribution of the size of the difference between the mean of each group at each timepoint (since shuffling the group labels eliminates any difference between the groups that might be present in the original data). Timepoints where the difference between the means of the real data were larger than 95% of the differences between the means of the shuffled data were considered to significantly differ between the groups. Following this, to control for the multiple comparisons across time, the duration of each significant period (periods showing consecutive timepoints of significant differences between the groups) in the real data were required to exceed 95% of the durations of the periods where the difference between means from a specific null distribution exceeded 95% of the other null distribution. Since neural activity from one timepoint to the next is highly correlated, we should expect real effects to be significant for longer than the effects from the null distribution, making this a physiologically appropriate method of multiple comparison control. This approach is commonly implemented in EEG research (Koenig et al. 2011, Bailey et al. 2019, Bailey et al. 2020, Habermann et al. 2018).

## 3. Results

### 3.1 N100 Amplitude

Figures 1 and 2 depict TEPs from single pulse stimulation to F3 and F4 respectively, and Figures 3 and 4 depict pp100-corrected TEPs from F3 and F4 respectively. Baseline comparisons of log transformed N100 amplitudes within our a priori selected N100 window between all three groups showed weak evidence for a main effect of group, which did not reach the threshold for statistical significance after controlling for multiple comparisons F(2,57) = 2.970, p = 0.059, η_p_^2^ = 0.094, FDR-p = 0.197 (Figure 5 and Table 1). No interaction was found between group and stimulation type (single or pp100) F(2,57) = 1.553, p = 0.220, FDR-p = 0.367, η_p_^2^ = 0.052, nor between group and hemisphere stimulated (right or left) F(2,57) = 0.372, p = 0.691, FDR-p = 0.819, η_p_^2^ = 0.013, nor three way interaction between group, stimulation type and hemisphere stimulated F(2,57) = 0.307, p = 0.737, FDR-p = 0.819, η_p_^2^ = 0.011.

**Figure 1.**
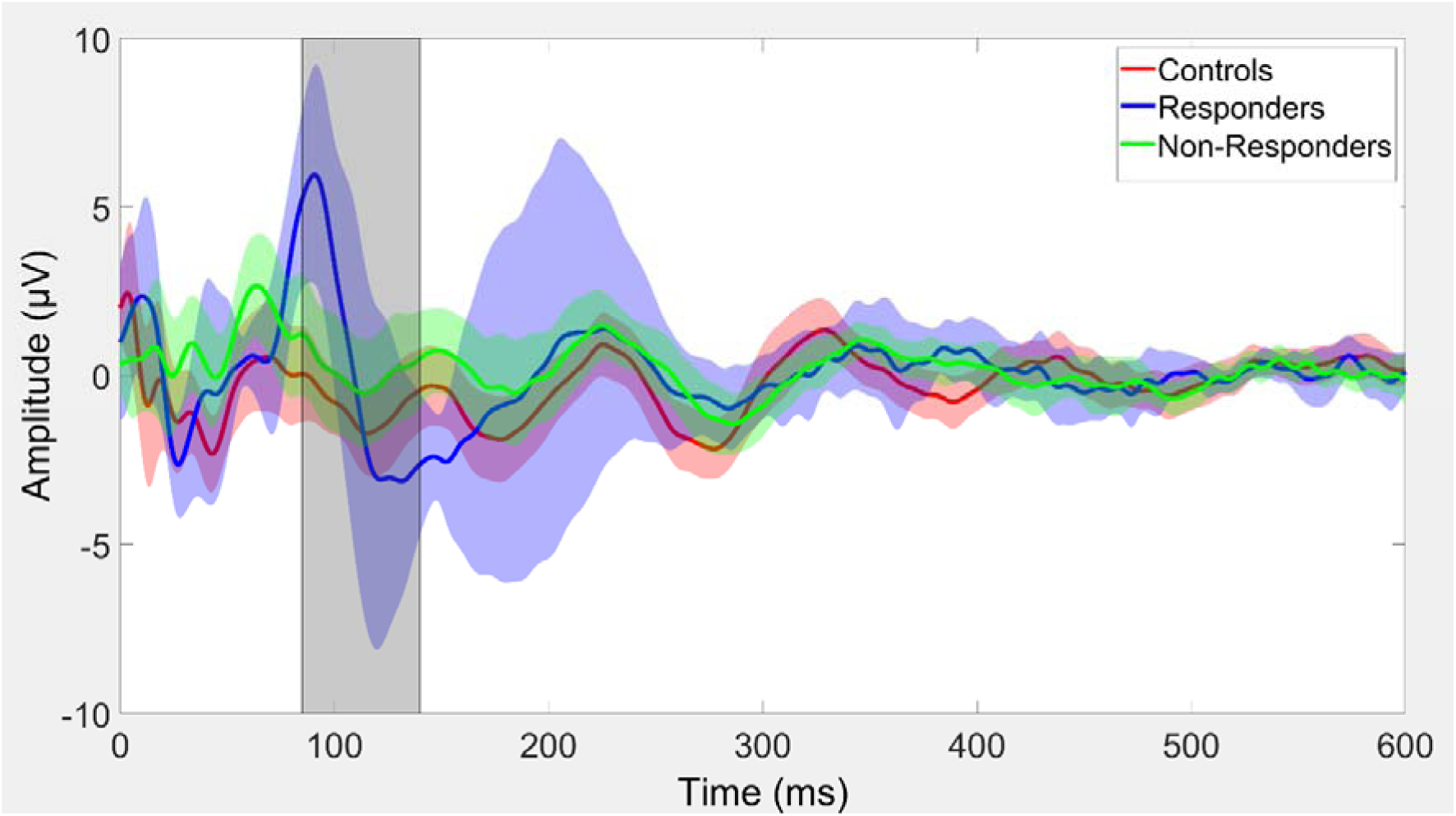
TEPs from F3 in response to single pulse LDLPFC stimulation from each group at BL. Error shading represents 95% confidence intervals, and the N100 window for analysis is shaded in grey.

**Figure 2.**
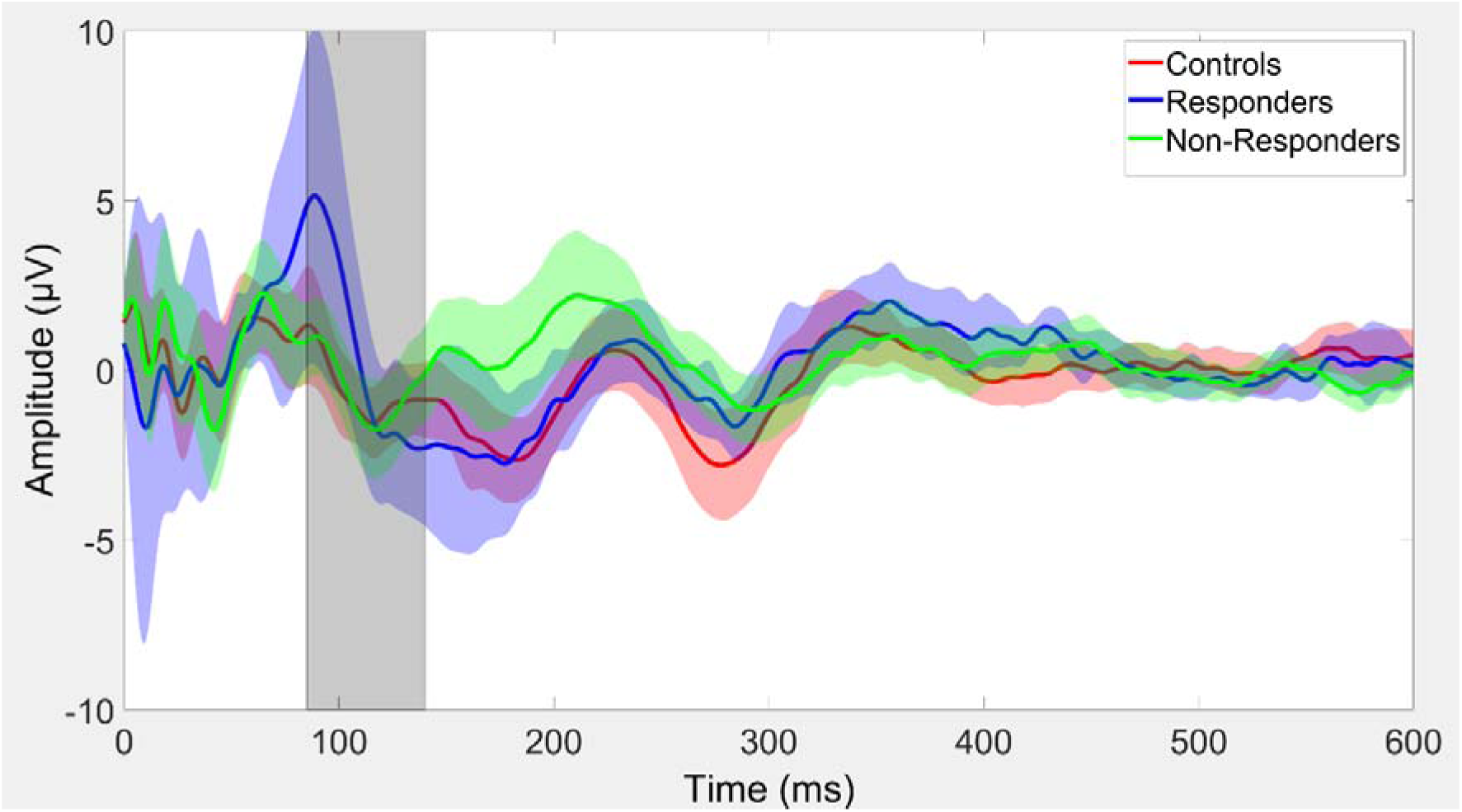
TEPs from F4 in response to single pulse RDLPFC stimulation from each group at BL. Error shading represents 95% confidence intervals, and the N100 window for analysis is shaded in grey.

**Figure 3.**
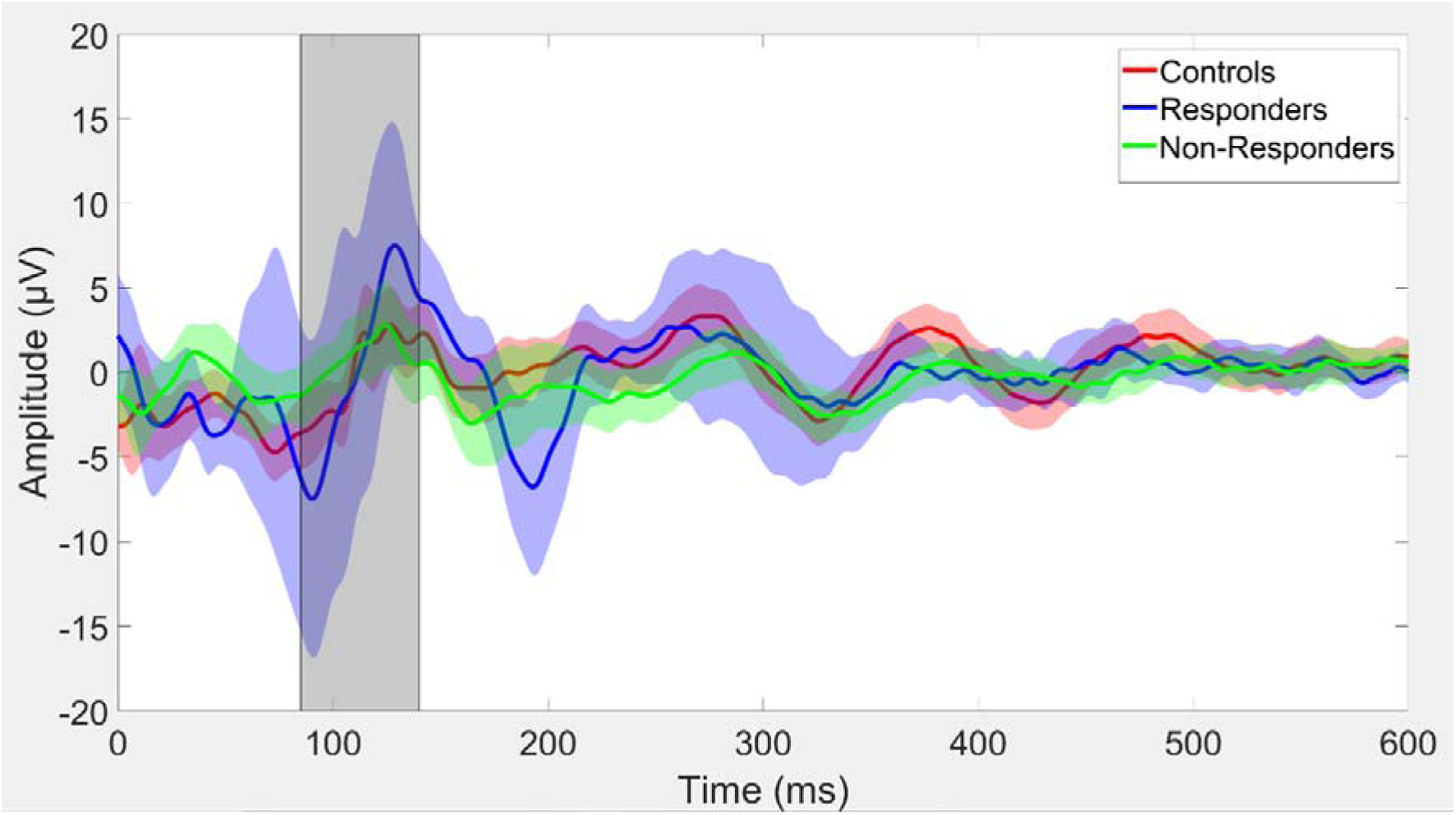
TEPs from F3 in response to pp-100 LDLPFC stimulation after correction for both the conditioning and test pulse from each group at BL. Error shading represents 95% confidence intervals, and the N100 window for analysis is shaded in grey.

**Figure 4.**
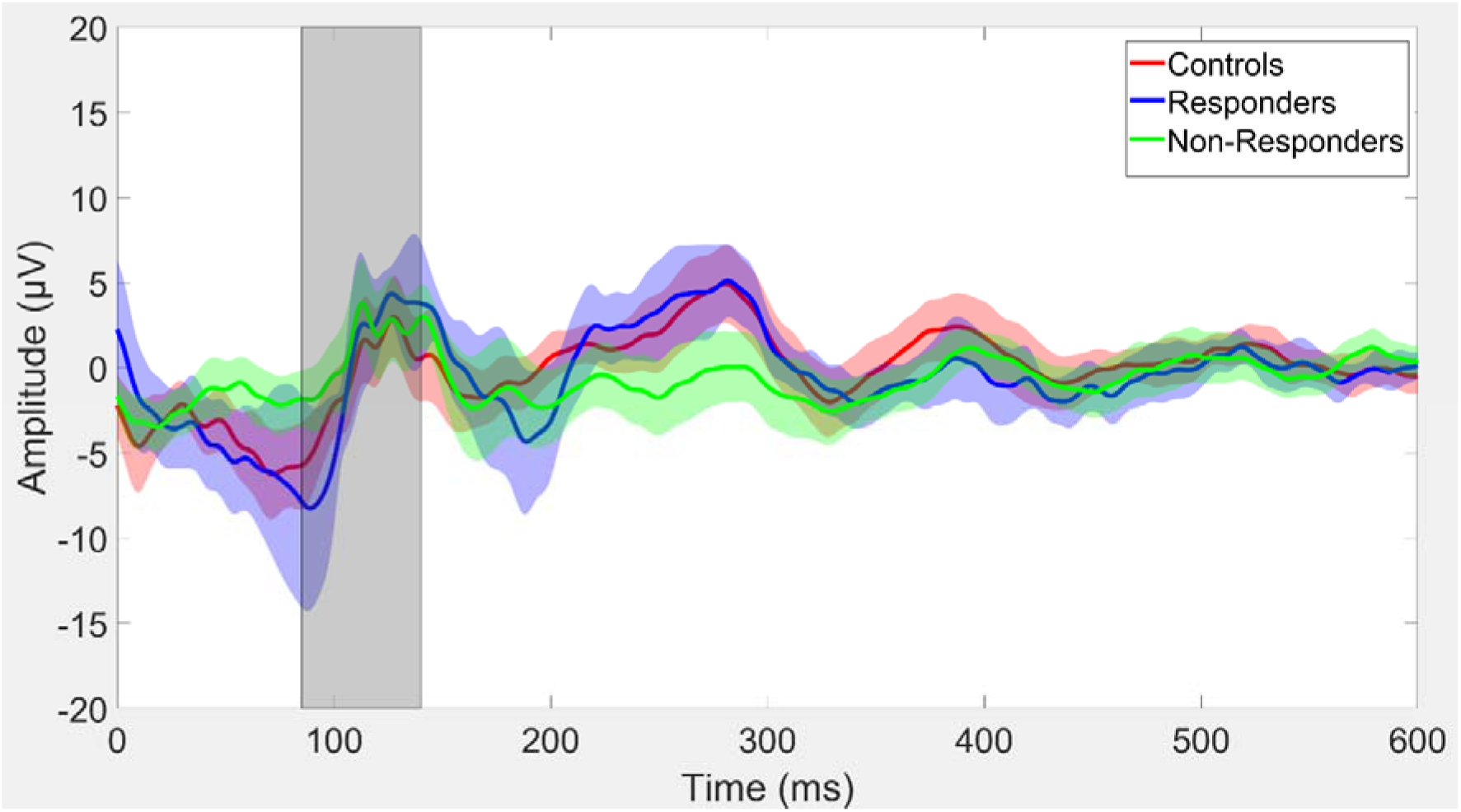
TEPs from F4 in response to pp-100 RDLPFC stimulation after correction for both the conditioning and test pulse from each group at BL. Error shading represents 95% confidence intervals, and the N100 window for analysis is shaded in grey.

**Figure 5.**
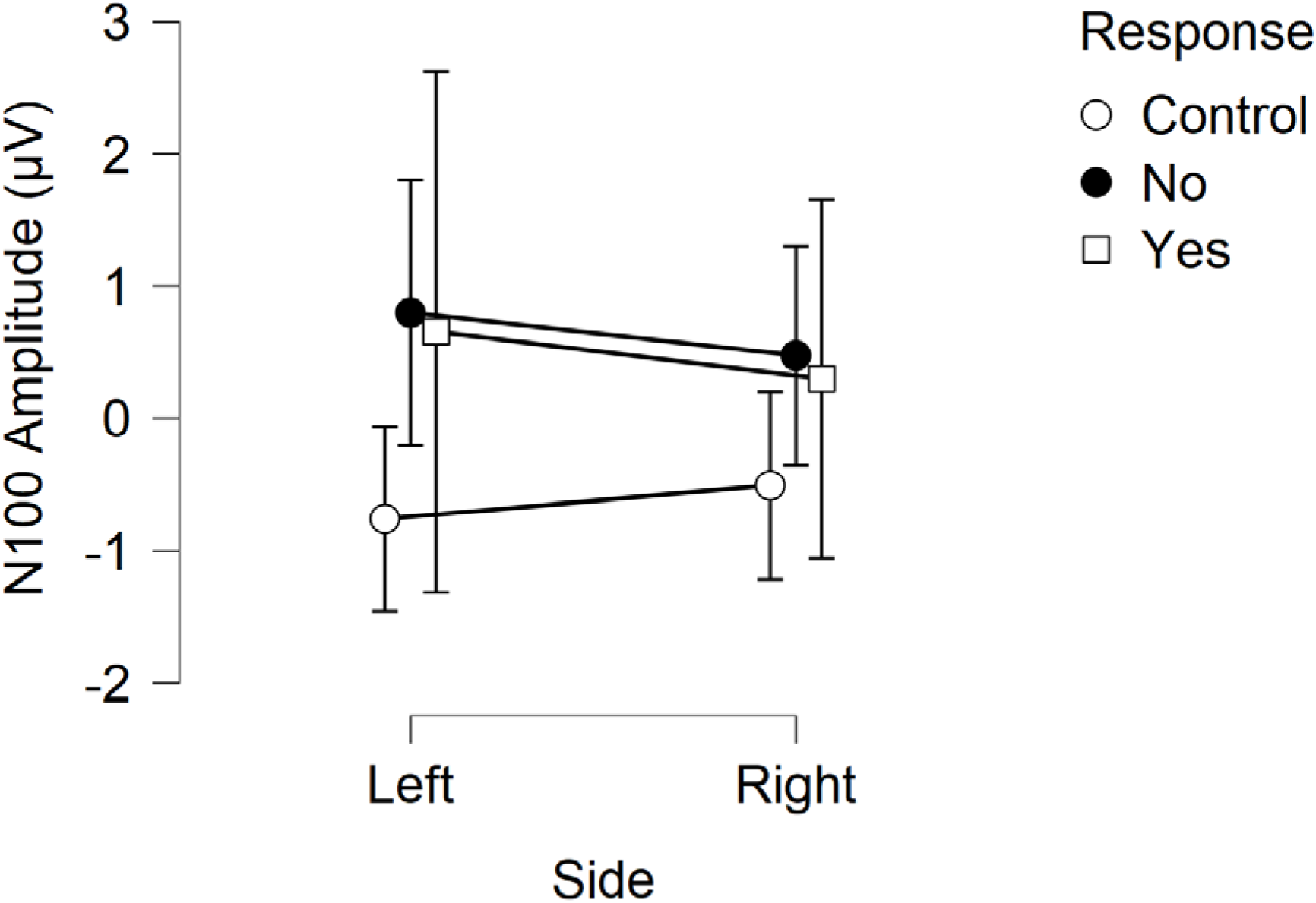
N100 amplitudes from each group, per side of stimulation. Error bars reflect 95% confidence intervals.

**Table 1.**
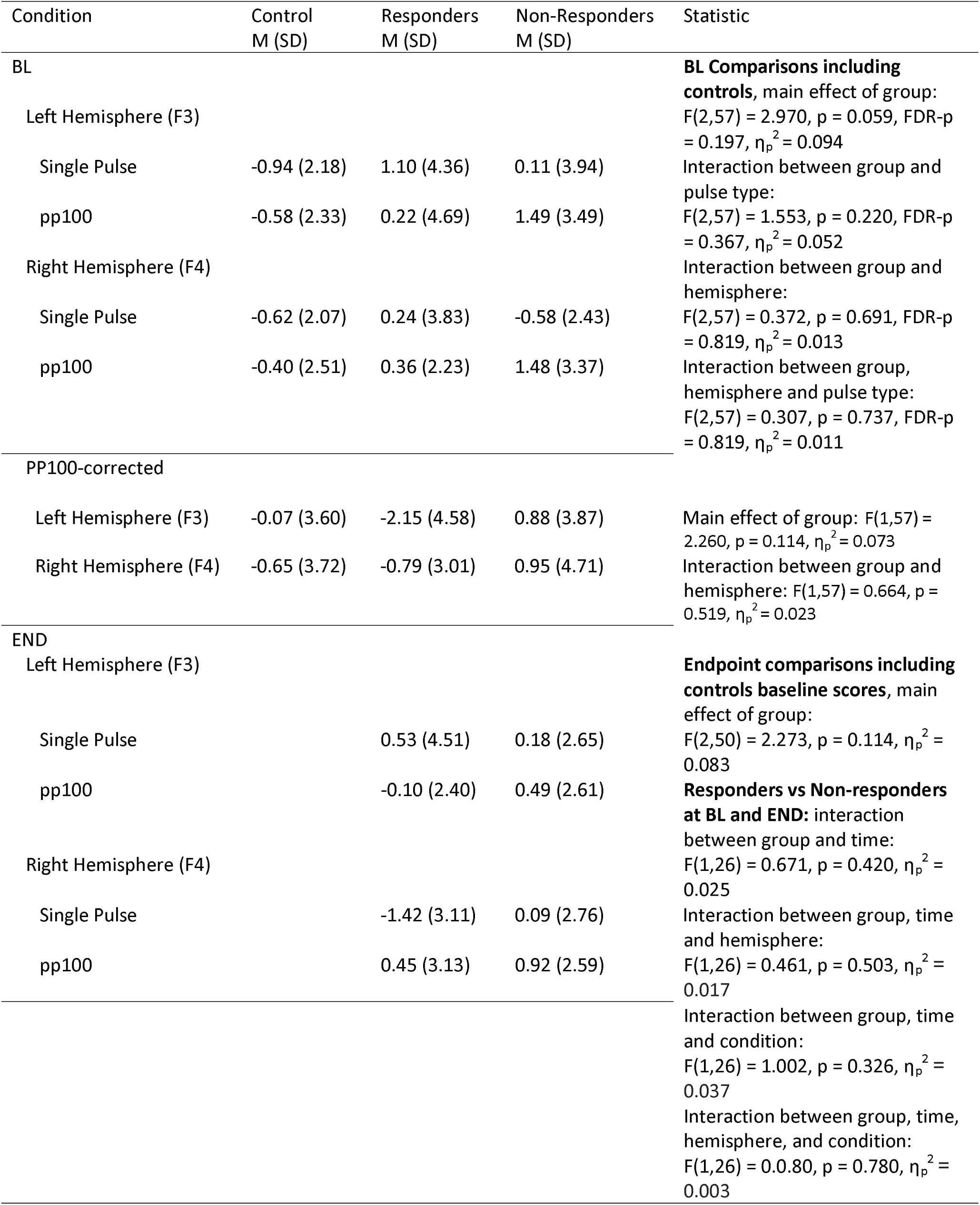
N100 Amplitude values and statistics

**Table 2.**
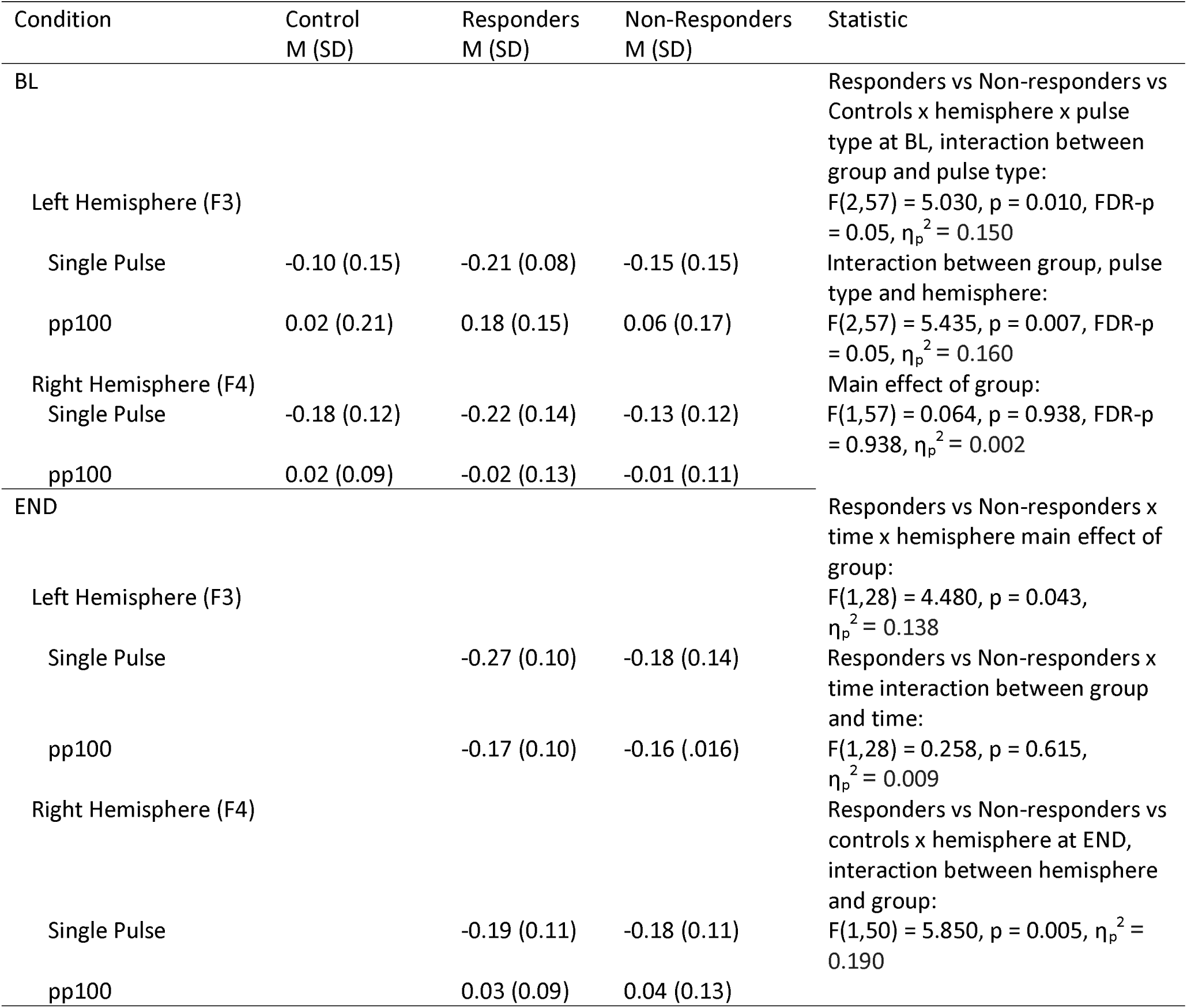
N100 Slope values and statistics

However, because the main effect of group approached significance, and inspection of the means within our a priori selected N100 window indicated this was due to the control group showing larger values than either of the depression groups, while the responder and non-responder groups showed almost identical means (see Figure 5), and given the small sample sizes, an exploratory repeated measures ANOVA was performed that combined the two depression subgroups. This analysis showed that the control group showed significantly more negative N100 amplitudes than the combined responder and non-responder depression group F(1,58) = 5.955, p = 0.018, η_p_^2^= 0.093. It is worth noting that the pattern of TEP response within the N100 window shown by the responder group differs from the other two groups, with a large and late P60 peak, and a large and late N100 peak (see Figures 1 and 2). Since the positive P60 peak is within the N100 window, the responder group showed a more positive mean N100 amplitude within our a priori selected N100 window than the control group, despite seeming to show a more negative N100 peak (which peaked later in the N100 window). We have performed an exploratory analysis without an a priori selected N100 window to assess this pattern, reported later in the results section.

With regards to the pp100-corrected comparisons, no significant main effect of group F(1,57) = 2.260, p = 0.114, η_p_^2^ = 0.073, or interactions between group and hemisphere were detected F(1,57) = 0.664, p = 0.519, η_p_^2^ = 0.023 (Figures 3 and 4).

Comparisons of N100 amplitudes at END (including baseline values from controls) showed no difference between groups F(2,50) = 2.273, p = 0.114, η_p_^2^ = 0.083. Neither did the end point comparisons show an interaction between group and stimulation type (single or pp100) F(2,50) = 0.084, p = 0.920, η_p_^2^ = 0.003, nor between group and hemisphere stimulated (right or left) F(2,50) = 0.676, p = 0.513, η_p_^2^ = 0.026, nor three way interaction between group, stimulation type and hemisphere stimulated F(2,50) = 1.546, p = 0.223, η_p_^2^ = 0.058. There was also no significant interaction between response group and timepoint F(1,26) = 0.671, p = 0.420, η_p_^2^ = 0.025. Nor were there any interactions between response group, timepoint and pulse type or hemisphere (all p > 0.10).

### 3.2 N100 Slope

Comparisons of N100 slopes showed no significant difference for group F(2,57) = 0.064, p = 0.938 FDR-p = 0.938, partial eta squared = 0.002. A significant interaction was found between group and stimulation type (single or pp100) F(2,57) = 5.030, p = 0.010, FDR-p = 0.05, partial eta squared = 0.150 (Figure 6), and between group, stimulation type, and hemisphere stimulated (right or left) F(2,57) = 5.435, p = 0.007, FDR-p = 0.05, partial eta squared = 0.160. There was no significant interaction between group and hemisphere stimulated F(2,57) = 0.978, p = 0.382, FDR-p = 0.546, partial eta squared = 0.033.

**Figure 6.**
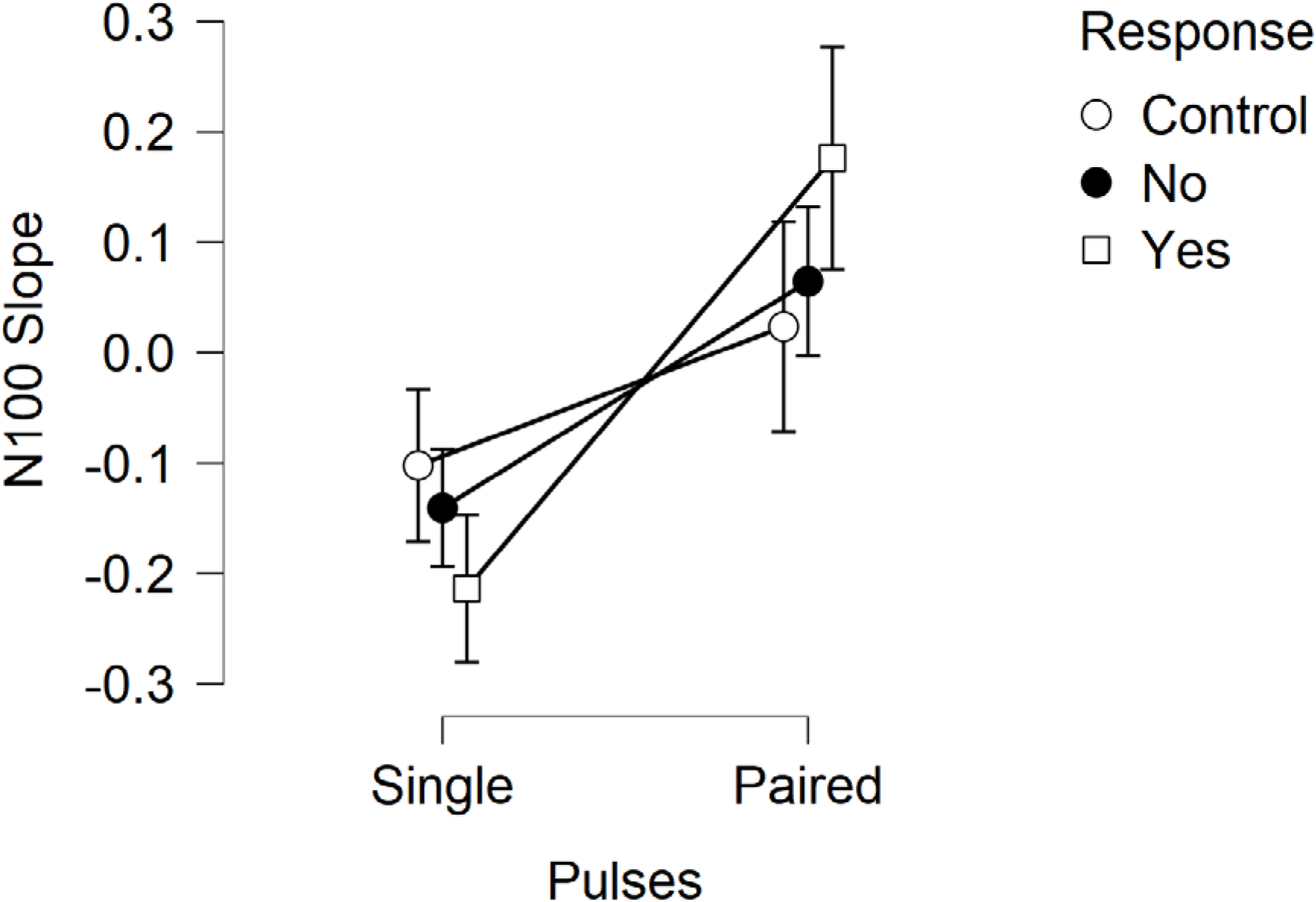
N100 slopes at F3 from each group in response to left sided stimulation. Error bars reflect 95% confidence intervals.

Parsing out the reason for the interactions by restricting comparisons to responders and non-responders only and to the left hemisphere only indicated a significant interaction between response group and stimulation type F(1,37) = 7.940, p = 0.008, partial eta squared = 0.177, with responders showing steeper negative slopes to single pulses (Figures 7) and steeper positive slopes to pp100 trials. However, between group comparisons restricted to single or paired pulse separately were non-significant when compared with a between groups t-test (single pulse t(37) = 1.602, p = 0.118, paired pulse t(37) = 1.867, p = 0.07), so the result was due to within group differences in the pattern of N100 slope to single and pp100 pulses. Responders showed negative slopes to single pulses (M = -0.21, SD = 0.10) and positive slopes to pp100 (M = 0.18, SD = 0.15), t(10) = 5.605, p < 0.001. Non-responders showed less steep negative slopes to single pulses (M = -0.14, SD = 0.14) and less steep positive slopes to pp100 (M = 0.06, SD = 0.17) t(27) = 6.64, p < 0.001. No significant main effect of group was present when including both single and pp100 in the same comparison F(1,37) = 0.215, p = 0.646, partial eta squared = 0.006.

**Figure 7.**
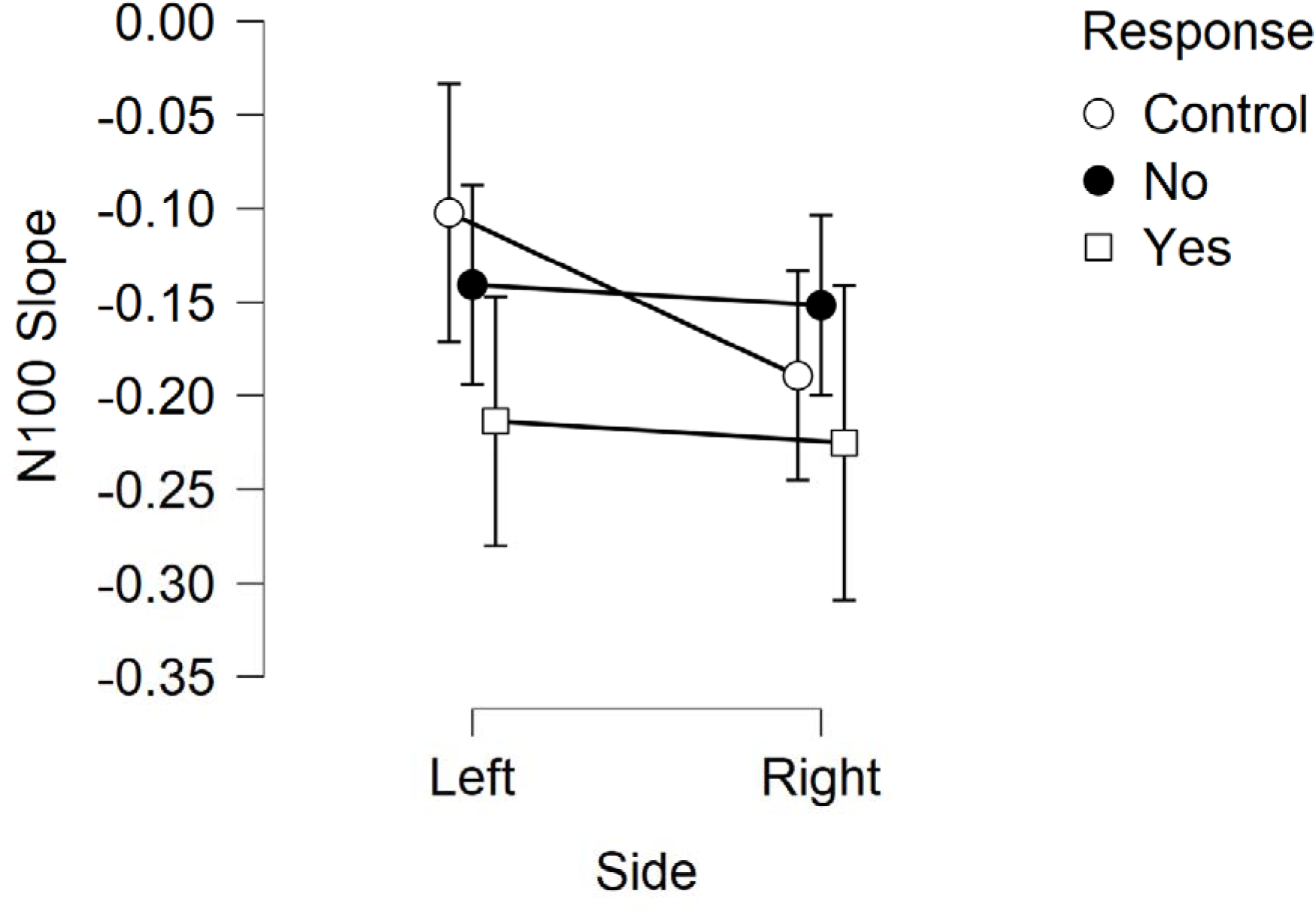
N100 slopes at F3 and F4 in response to single pulse left hemisphere stimulation and right hemisphere stimulation respectively from each group in response. Error bars reflect 95% confidence intervals.

In contrast, comparisons between responders and non-responders restricted to the right hemisphere only indicated no significant interaction F(1,38) = 1.345, p = 0.253, partial eta squared = 0.034, nor main effect of group F(1,38) = 1.709, p = 0.199, partial eta squared = 0.043.

When comparisons were made between responders and non-responders and between hemispheres for single pulse stimulations only, there was no significant interaction between hemisphere and group F(1,37) = 0.000, p = 0.992, partial eta squared = 0.000, but a significant main effect of group was present F(1,37) = 4.874, p = 0.034, partial eta squared = 0.116, with responders showing more negative slopes than non-responders in response to stimulation at both hemispheres (Figure 8).

**Figure 8.**
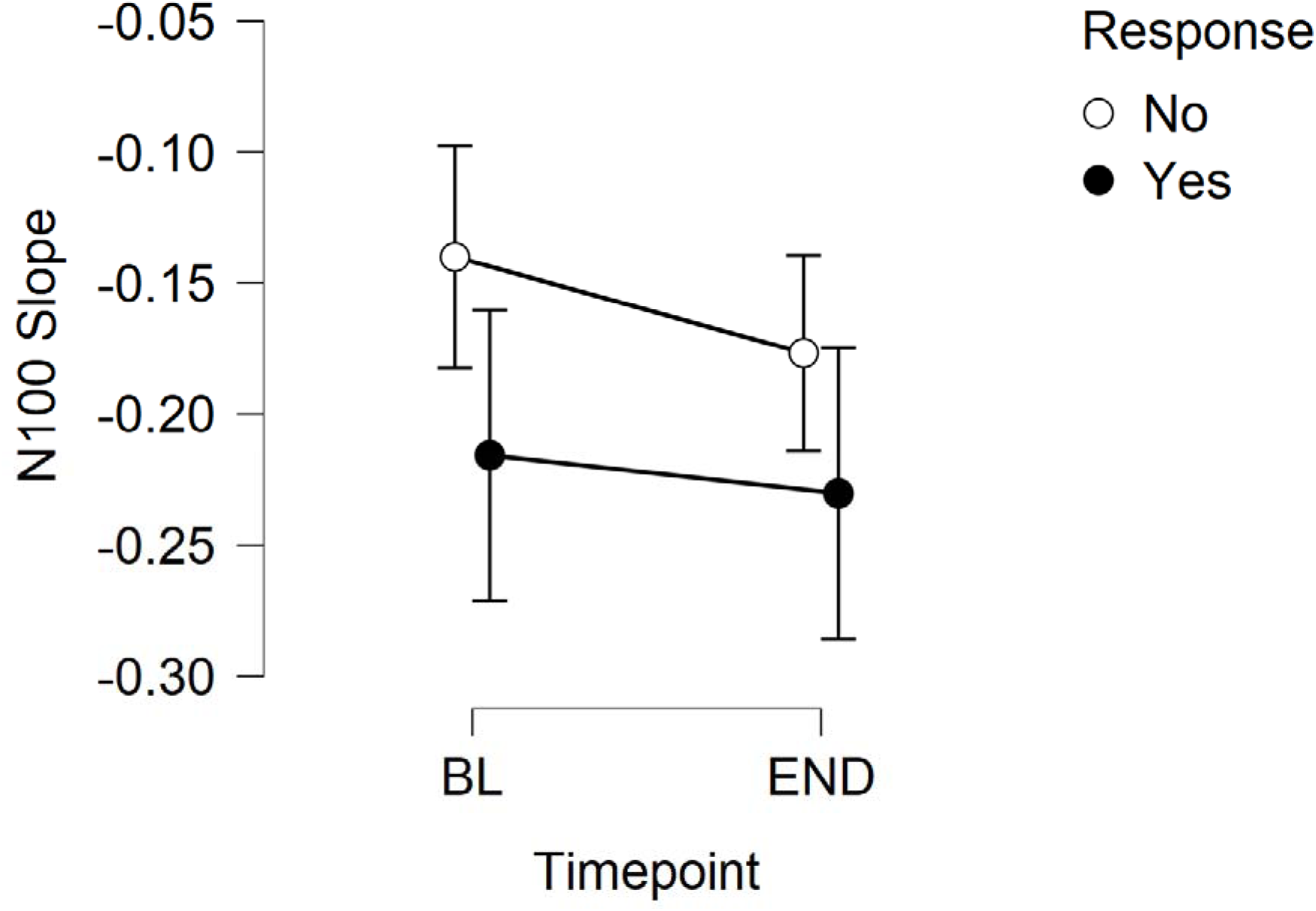
N100 slopes at baseline and end from each group in response to single pulse stimulation, averaged across both hemispheres. Error bars reflect 95% confidence intervals.

In contrast, when comparisons were made between responders and non-responders and between hemispheres for paired pulse stimulations only, there was no significant interaction between hemisphere and response F(1,37) = 3.432, p = 0.072, partial eta squared = 0.085, and no significant main effect of group was present F(1,37) = 1.621, p = 0.211, partial eta squared = 0.042.

To summarise, we found differences between responders and non-responders in the pattern of response to single pulse stimulation vs pp100 stimulation. The interaction between group, hemisphere and stimulation type may have been driven by a more positive slope in response to paired pulses on the left side in responders compared to non-responders, but when the interaction was explored the result was only significant at a trend level. An exploratory Receiver Operator Characteristic (ROC) curve analysis was used to assess the ability of this difference between left hemisphere single and paired pulse slopes from each individual to predict treatment response. A linear discriminant was fitted over the distribution of the data with MATLAB’s ‘fitcdiscr’ function. The ROC curve was used to provide an estimate of the model’s optimal sensitivity, specificity and accuracy thresholds. This analysis provided a sensitivity value of 0.455, a specificity of 1.00, accuracy of 0.846, and area under the curve of 0.737 (Figure 9).

**Figure 9.**
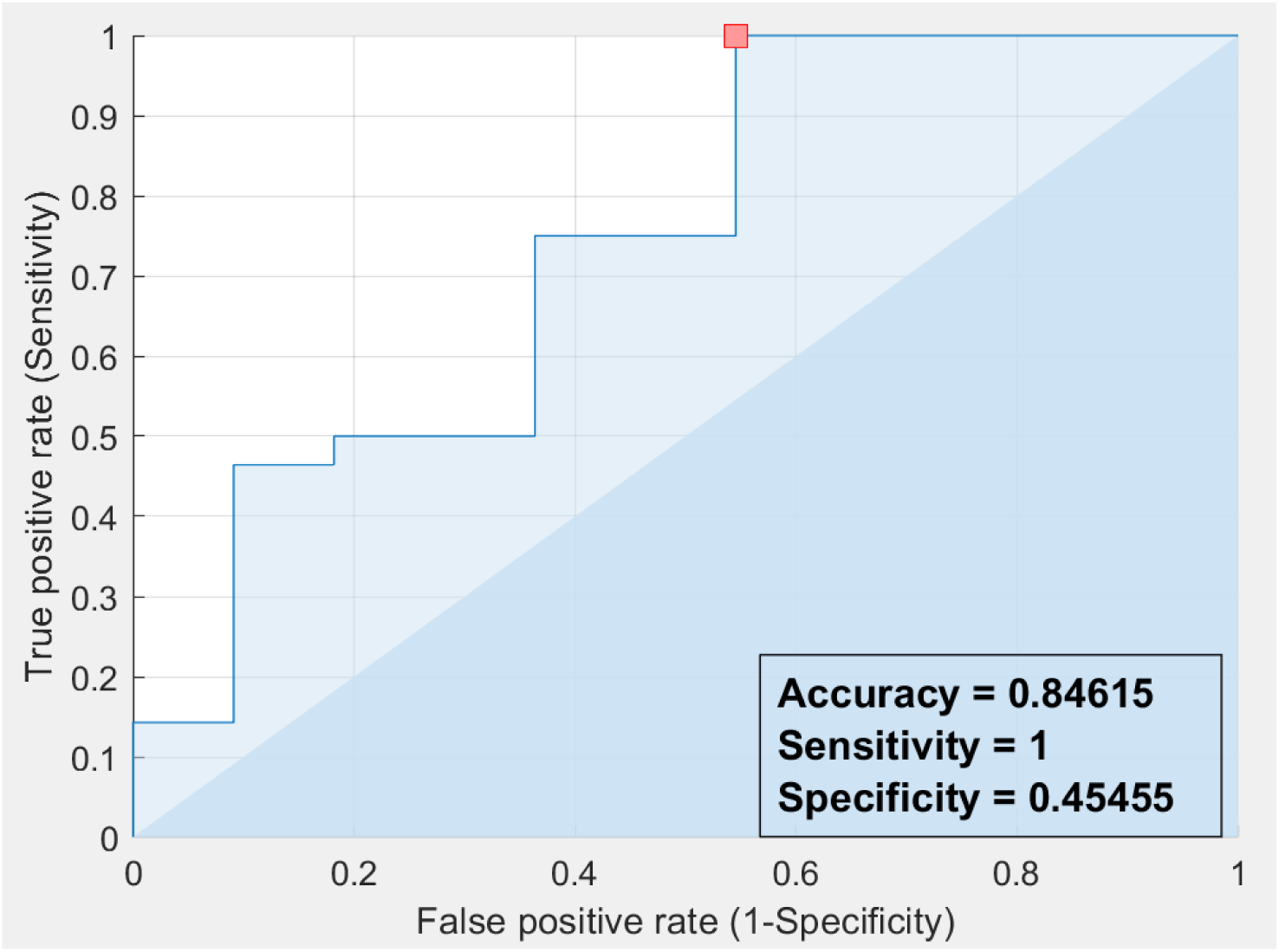
A linear Receiver Operator Characteristic (ROC) curve analysis of the ability of the difference between left hemisphere single pulse and paired pulse N100 slopes to predict response to rTMS treatment. The red square indicates the optimal thresholding point.

Given that it appeared that the responders showed steeper negative N100 slopes than both non-responders and controls, an exploratory test including BL and END timepoints was conducted to determine whether this result reflected an abnormal marker that is normalised by successful rTMS treatment, or whether the steep negative N100 slope in responders is a trait that may confer susceptibility to the effects of rTMS treatment leading to treatment success (Figure 9). This was comprised of a repeated measures ANOVA comparing responders and non-responders in the single pulse TMS with both hemispheres included, at both BL and END timepoints. The results showed a main effect of group F(1,28) = 4.480, p = 0.043, η_p_^2^ = 0.138, with responders still showing more negative N100 slopes than non-responders. No significant interaction was present for the time by group comparison F(1,28) = 0.258, p = 0.615, η_p_^2^ = 0.009 or time by group by hemisphere comparison F(1,28) = 2.452, p = 0.129, η_p_^2^ = 0.081.

Additionally, an exploratory comparison of responder and non-responder END single pulse measures to control BL measures (since controls only provided data at BL) (repeated measures ANOVA, 3 groups x 2 hemispheres) did not show a main effect of group F(2,50) = 2.044, p = 0.140, η_p_^2^ = 0.076, but did show a significant interaction between group and hemisphere F(2,50) = 5.850, p = 0.005, η_p_^2^ = 0.190. A post-hoc Tukey test restricted to the left hemisphere showed responders had more negative N100 slopes than controls (p = 0.006) while responders and non-responders did not differ (p = 0.200) and controls and non-responders did not differ (p = 0.111).

Visual inspection of TEP waveforms (Figure 1) suggested that the steeper N100 slopes in the responder group in response to single pulse TMS to the LDLPFC at baseline might have been due to a large late P60 amplitude and large late N100 amplitude in the responder group. Performing statistical comparisons of windows of TEP activity based on visual inspection of the windows showing the largest difference is not valid (Kilner, 2013), so statistical comparisons of the specific P60 and N100 time windows where the differences seemed apparent were not performed. However, permutation statistical comparisons of the single pulse condition applied to the LDLPFC across the entire epoch showed a significant difference between responders and non-responders from 128 to 160ms (32ms in duration), which lasted longer than 95% of the significant periods from the null distributions (27ms). This difference was due to the responder group showing a larger (more negative) amplitude during this late N100 period (p = 0.0141 averaged within the significant period, Cohen’s d = 1.033). Additionally, there was a significant difference between the responder and control group from 80 to 105ms (25ms in duration) which lasted longer than 93% of the significant periods from the null distributions (the 95% threshold was 27ms). This difference was due to the responder group showing a larger (more positive) amplitude during this late P60 period than the control group (p = 0.0058 averaged within the significant period, Cohen’s d = 1.340). When data was averaged within this period, the responder group also showed significantly larger amplitudes than the non-responder group (p = 0.0293, Cohen’s d = 0.843).

To assess the consistency of this finding across participants, a violin plot was produced for the mean amplitude within the significant windows (from 85 to 105 ms and 128 to 160ms, see Figure 10). This plot showed that only 3/12 responders had smaller late P60 amplitudes or smaller N100 amplitudes than the non-responder or control median at baseline. Together, these results suggest that the steeper negative N100 slopes (perhaps driven by more positive and late P60 amplitudes and more negative late N100 amplitudes) in responders reflect a trait characteristic that is not modified by successful rTMS treatment, but may be a trait related to successful treatment response.

**Figure 10.**
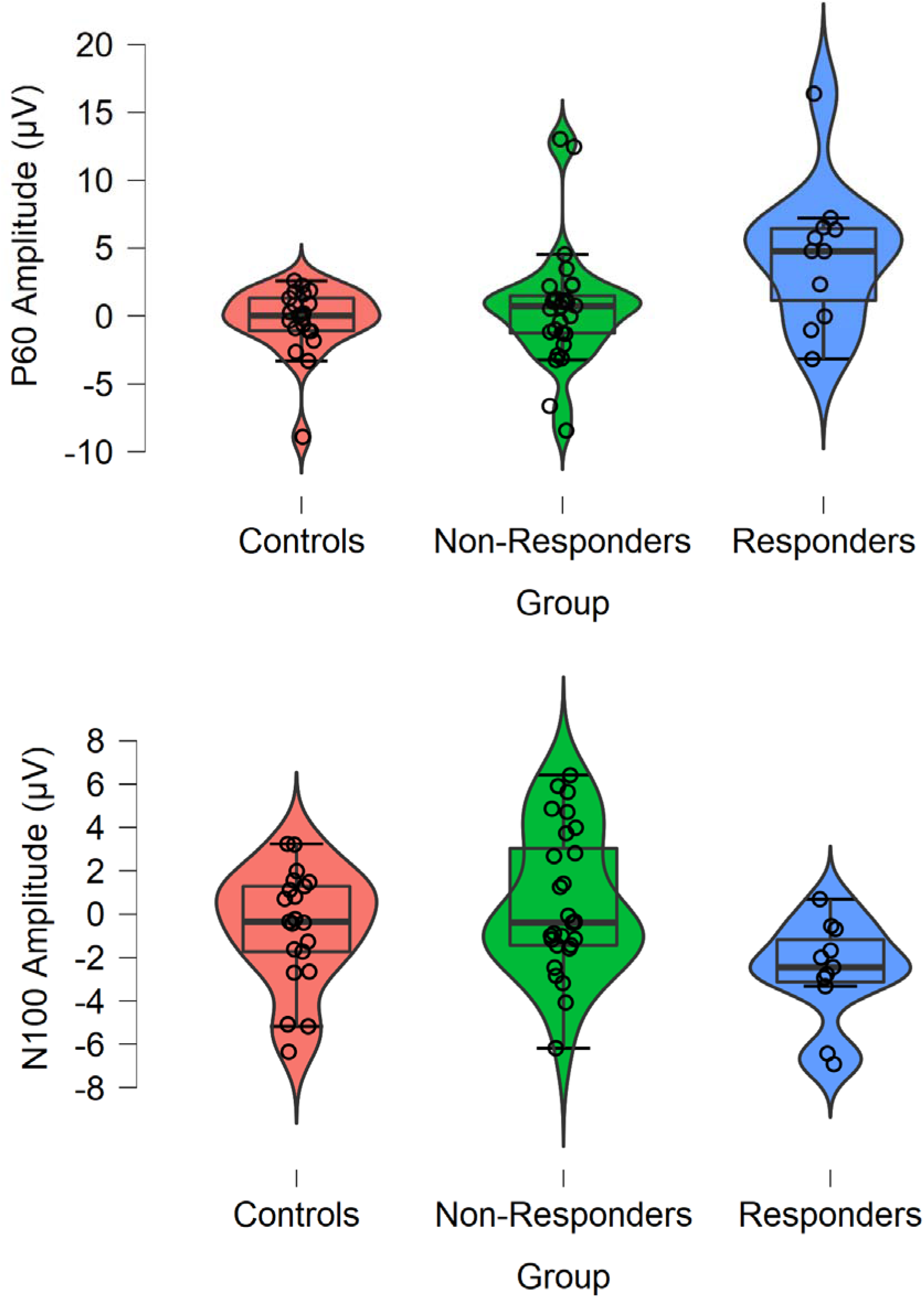
Violin plot depicting P60 amplitudes (top) and N100 amplitudes (bottom) from the significant windows from permutation testing (from 85 to 105 ms and 128 to 160ms respectively) in response to single pulse stimulation to the LDLPFC.

### 3.3 Theta power

No significant main effect or interaction with response group was detected in theta power (all p > 0.20, results can be viewed in table 3).

**Table 3.** Theta BL Corrected power values and statistics. Note that comparisons were restricted to single pulses only.

| Condition | Control<br>M (SD) | Responders<br>M (SD) | Non-Responders<br>M (SD) | Statistic |
| --- | --- | --- | --- | --- |
| Left Hemisphere<br>Single Pulse<br>F3 | 1.76 (0.81) | 2.15 (1.30) | 1.61 (0.42) | Main effect of group:<br>$F(2,57) = 1.633$, $p = 0.204$, $FDR-p = 0.367$, $\eta_p^2 = 0.054$<br>Group by Hemisphere interaction:<br>$F(2,57) = 1.825$, $p = 0.170$, $FDR-p = 0.367$, $\eta_p^2 = 0.060$ |
| Right Hemisphere<br>Single Pulse<br>F4 | 1.47 (0.33) | 1.77 (0.65) | 1.67 (0.62) |  |

## 4. Discussion

This study examined whether TMS-EEG measures of neural activity could differentiate responders and non-responders to rTMS treatment for depression. The results showed that N100 amplitude did not differ between responders and non-responders, but the healthy control group showed more negative N100 amplitudes than the combined depression group. The N100 slope on the other hand, did differentiate responders and non-responders, with responders showing a steeper negative slope for single pulses and steeper positive slope following pp100 at F3 in response to stimulation to the left hemisphere (slopes were negative in response to single pulses, and positive in response to pp100 pulses, so responders showed steeper slopes in both directions, which resulted in a significant interaction between stimulation type and response group in the left hemisphere). Exploration of this finding suggested that the difference might have been driven by responders showing large and late P60 amplitudes in the left hemisphere, so that the slope from the positive late P60 to the negative N100 was largest in the responder group. However, differences in the late P60 amplitude were not an a priori hypothesis, so this result should be considered exploratory. An exploratory ROC analysis of the difference between the left hemisphere single and paired pulse N100 slope also indicated excellent sensitivity (although poor specificity), suggesting the slope difference may be helpful in predicting non-response to rTMS for depression. Theta power in response to TMS pulses did not differentiate any of the groups.

The brain’s response to single pulse TMS is emerging as an important biomarker for differentiating people with depression from healthy individuals. Voineskos and colleagues (2019) found that people with depression showed larger N45, P60 and N100 TEPs than healthy individuals following stimulation of left DLPFC, Dhami and colleagues (2020) found increased N100 and P200 amplitudes in youth with depression, whereas Hill and colleagues (2021) found increased TMS-evoked oscillations in the delta (1-3 Hz), theta and alpha (8-12 Hz)-bands. Our results partially align with these findings, showing larger N100 amplitudes and steeper N100 slopes at baseline in people who responded to rTMS treatment compared to those who did not respond, and also compared to healthy individuals. We also showed preliminary evidence of a larger P60 amplitude in responders.

TEPs also appear sensitive to changes in neural states induced by brain stimulation-based therapies. Following rTMS treatment for depression, Voineskos and colleagues found that the N45 and N100 TEPs from left DLPFC decreased in amplitude with change in N100 correlating with treatment response, suggesting treatment may partially return TEP amplitudes towards healthy levels. Following convulsive therapies, Sun and colleagues (2016) showed baseline N100 amplitude and suppression of TEPs with pp100 predicted reduction in suicidal symptoms following magnetic seizure therapy (MST) and that TEPs increased in amplitude following MST (Sun et al 2018). Casarotto and colleagues (2013) showed increases in the amplitude and slope of the P25 and N45 following electroconvulsive therapy (ECT), whereas Hill and colleagues (2021) found decreases in TMS-evoked delta and theta-band oscillations following both MST and ECT. In contrast to these findings, we did not find any evidence of changes in TEP characteristics following rTMS in either responders or non-responders to rTMS treatment. While the means from each group suggested that responders showed an increase in N100 amplitudes from baseline to treatment end, such that their N100 values were more similar (and not significantly different from the control participants at this timepoint), there was also no significant interaction between response group and timepoint. Of note, the participants in this study underwent a combination of different rTMS therapies, including left sided high frequency, right sided low frequency and sequential left and right sided treatment. It is possible that the combination of treatments across our group may have masked changes specific to any individual treatment regime. Future studies with larger samples for each individual treatment type are required to better characterise how rTMS alter TMS-evoked brain activity.

The N100 TEP is likely to reflect a combination of neural mechanisms, including TMS-evoked neural activity possibly related to cortical inhibition, but also sensory activity related to clicking noise of the TMS pulse and the scalp sensation caused by TMS. Cortical inhibition and associated biomarkers are the subject of interest in a variety of neurological and psychiatric contexts. In particular, GABAb receptor-mediated inhibition has been implicated in cortical functioning (Kohl & Paulsen, 2010), with dysfunctional GABAb receptor-mediated inhibition implicated in epilepsy (Schuler et al., 2001) and schizophrenia (Rogasch, Daskalakis, & Fitzgerald, 2014). Correlates of GABAb receptor-mediated cortical inhibition such as the N100 peak have also found to have been reduced in children with ADHD (Bruckmann et al., 2012; D’Agati et al., 2013). The GABAb receptor has also been implicated in depression, both in human studies (Romeo et al. 2018) and with GABAb receptor knockouts having been associated with increased depression like behaviour (Mombereau et al., 2004), while receptors agonists such as Baclofen showing antidepressant-like effects in rats (Felice et al., 2020; Khan et al., 2016). Baclofen increases N100 peak amplitude over motor cortex in healthy individuals (Premoli et al., 2014;), and N100 has been implicated in both transient and persisting measures of post-synaptic inhibition showing relationships to the cortical silent period (Farzan et al., 2013) and pp100 (Rogasch et al., 2013), respectively. The difference in N100 amplitude between the depression and control groups in the current study may therefore reflect altered GABAb receptor-mediated inhibitory function. Given these findings, one possible cause of changes in the N100 peak is changes in GABAb-receptor mediated cortical inhibition in people with depression. However, it is worth noting that our analysis of the pp100-corrected N100 showed no differences between the groups, suggesting the amount of N100 suppression produced by the pp100 conditioning pulse did not differentiate the three groups.

It is also important to note that the nature of the N100 has been contested amongst more recent literature. Due to the manner in which EEG signals reflect a contribution from both excitatory and inhibitory activity, it is difficult to clearly understand what the N100 represents. Research has argued that both the auditory and muscular activity arising from TMS are larger in amplitude and last for longer than any cortical neural activity, therefore hiding or confounding any contribution from pulse related inhibition or excitation in TEPs (Conde et al., 2019; Korhonen et al., 2011; Rogasch et al. 2014). Moreover, auditory activation appears to be time-locked to the N100 TEP in particular (Nikouline, Ruohonen & Ilmoniemi, 1999; Rogasch et al. 2014; ter Braack, de Vos & van Putten, 2013, Biabani et al. 2019, Biabani et al. 2021, Fernandez et al. 2021). A study by Conde et al., (2019) also found that simulations of TMS coil clicks delivered to common TMS target sites produced similar patterns of EEG activity when compared to active TMS delivery. Additionally, removing auditory artefacts during data analysis has been shown to lead to significant reductions in EEG amplitudes (Rogasch et al. 2014, Biabani et al. 2019). This has led to suggestions that the N100 may largely reflect sensory-related brain activity (ter Braack et al., 2013; Conde et al., 2019). The current study did use a sound-masking protocol. Sound-masking protocols have been associated with less distortion of cortical EEG activity (Rogasch et al. 2014; ter Braack et al., 2013), however residual sensory activity is often present regardless of auditory masking (Gordon et al. 2018, Conde et al. 2019, Biabani et al. 2019, Biabani et al. 2021, Fernandez et al. 2021). Given these findings, it is also possible that differences in the N100 observed in this study could reflect differences in sensory processing of the TMS pulse between groups. Despite this, the N100 did still differ between the groups, and the ROC analysis indicated that this difference offered predictive potential. As such, future research is recommended to determine the physiological underpinnings of this difference.

The lack of difference in theta activity is interesting when placed in comparison to our previous research examining EEG activity during the Sternberg task in the same group of participants (which did demonstrate a difference in fronto-midline theta activity between the responders and non-responders). The contrast between that result and the current study suggests that the differences between responders and non-responders in fronto-midline theta might be specifically elicited by the performance of cognitive processes. Cognitive processes have been shown to elicit fronto-midline theta, which is thought to reflect cognitive control, executive functioning, or attentional processes (Cavanagh and Shackman, 2015). The lack of a difference in theta oscillatory activity in the TMS-EEG results might suggest that the higher level of fronto-midline theta in responders might be endogenously produced in response to task demands rather than being an intrinsic resonance property of the network that can be stimulated by TMS to the DLPFC.

### 4.1 Limitations

There are a number of caveats and limitations to the current research. In terms of caveats, it should be noted that the effect size of the difference between responders and non-responders in N100 slope are moderate, and smaller than the effect size for other variables that have been shown to differentiate these two groups (Bailey et al. 2018, 2019). In particular, early response to rTMS treatment showed much larger effect sizes, so is likely to be a more accurate predictor (Bailey et al. 2018, 2019, Feffer et al. 2018). Alone, the N100 slope is unlikely to be a very accurate predictor, so would perhaps be best used in combination with other predictive variables to allow higher accuracy than individual variables used in isolation. Additionally, the sample size included in the current study is relatively small, particularly for the responder group. As such, it may be that the participants from the current study reflect a specific subsample, and that the results of analyses performed on this group may not generalise to the broader population of individuals with depression undergoing rTMS treatment. This small sample size could be a potential explanation for some of our null results. Future research should examine N100 slope in particular with larger sample sizes for comparison between responders and non-responders. Due to these low numbers, it is also possible that a sampling bias was present in our study, and this sampling bias may be responsible for the difference between the responder and non-responder groups. The small sample size also prevented separate analyses of the different treatment protocols involved in the study, and the different treatment protocols implemented are a limitation that increased the heterogeneity of the sample, so the results of the study can only speak to ‘response to rTMS treatment in general’. Future research with larger sample sizes should attempt to determine whether N100 slope measures might predict response to a particular treatment type. The single TMS-EEG timepoint obtained for the control group is another important limitation, which prevented the use of a repeated measures comparison including the control group to determine whether the MDD and control groups differed in changes in EEG response to TMS over time.

Additionally, the distribution of electrodes used in our EEG cap for this study was sparse, and a smaller number than some studies was used. As such, cluster-based statistics were not appropriate for analysis of our data. Future research should use more electrodes to provide a more rigorous and complete characterisation of the effect of TMS pulses on EEG data in responders and non-responders. Lastly, more research is required to determine the reason for the difference in N100 slope detected by the current study. Since responders showed N100 slopes that were different from both non-responders and controls, the results may suggest that the steeper N100 slope in the responder group reflects a particular underlying pathophysiology that is responsive to rTMS. However, because the difference in the responder group was still present at the endpoint, the results suggest that while rTMS did improve the group’s depression severity, it did not do so by resolving the pathophysiology.

### 4.2 Conclusions

Overall, the current results suggest that left hemisphere N100 slope, or perhaps the ratio of late P60 to N100 peaks in response to TMS stimulation to the left DLPFC might be able to differentiate responders and non-responders to rTMS treatment for depression at baseline. The ROC analysis suggested the within-individual differences between the single and pp100 N100 slopes provide excellent sensitivity, indicating these values may be useful for prediction of non-response to rTMS for depression. The consistency between baseline and endpoint differences suggests that this reflects a trait characteristic of the responder group rather than a dysfunctional mechanism that is resolved or improved by the rTMS treatment. As such, it may reflect a characteristic that positively interacts with the rTMS treatment, perhaps enabling increased plasticity or spreading of network effects related to the rTMS treatment. However, due to the limitations of the current study, future research with larger sample sizes is recommended to determine if our results replicate before they could be assessed for potential clinical applications.

## Registration

This study was registered with the Australian New Zealand Clinical Trials Registry (ACTRN12610000998044).

## Data Availability

We do not have ethical approval to share the data involved in this study.

